# Delays in COVID-19 Diagnosis and Hospitalization and Outcomes — New York City, New York, USA, October 2020–November 2021

**DOI:** 10.1101/2022.06.02.22275918

**Authors:** Laura E. Graf, Eric R. Peterson, Jennifer Baumgartner, Anne Fine, Corinne N. Thompson, Kathleen Blaney, Sharon K. Greene

## Abstract

COVID-19 patients diagnosed ≥3 days after symptom onset had increased odds of hospitalization. The 75^th^ percentile for diagnosis delay was 5 days for residents of low-privilege areas and Black and Hispanic people diagnosed before SARS-CoV-2 Delta predominance, compared with 4 days for other patients, indicating inequities in prompt diagnosis.

---

Delays in testing or care-seeking for certain respiratory infections can impede access to potentially life-saving treatments (*1, 2*), contributing to worse outcomes, including hospitalization and death (*3*). Reasons for delayed care-seeking include prohibitive costs, lack of access to healthcare services, and stigmatization (*4, 5*). However, little is known about the impact of care-seeking delays on COVID-19 outcomes in the United States (*6, 7*). We assessed relationships between delays in diagnosis of and hospitalization with COVID-19 and severe outcomes.

## The Study

The study population included NYC residents symptomatic with COVID-19-like illness and diagnosed with COVID-19 via a positive SARS-CoV-2 molecular or antigen test or hospitalized after exposure to a confirmed COVID-19 case during October 1, 2020**–**November 30, 2021. We hypothesized that increased time between symptom onset and diagnosis or hospitalization was associated with worse outcomes; accordingly, our exposures were time to diagnosis (days from symptom onset to diagnosis) and time to hospitalization (days from symptom onset to hospital admission), each classified into quintiles. COVID-19 symptoms and symptom onset date were collected through case investigation. Diagnosis date was defined as the earlier of specimen collection date of the first positive test or hospital admission date. Patients with an onset date following their diagnosis or admission date were excluded, effectively excluding healthcare-associated cases. Patients with a diagnosis or admission date >21 days after onset were excluded because true long delays could not be distinguished from data entry errors.

Our outcomes of interest were hospitalization, death, and length of hospital stay. A COVID-19 hospitalization was defined as a hospital admission +/- 14 days from a positive SARS-CoV-2 molecular or antigen test or hospitalization at the time of death from COVID-19. A COVID-19 death was defined as 1) occurring within +/- 30 days of a positive COVID-19 molecular test and not from external causes or 2) a cause of death of COVID-19 on the death certificate and a positive COVID-19 molecular test. Length of stay was defined as the number of days from hospital admission to discharge and was dichotomized at the median (>5 vs. ≤5 days) (Appendix: Statistical Analysis). The NYC Department of Health and Mental Hygiene’s Institutional Review Board determined this work was exempt human subjects research for which consent was not required.

Of 864,880 COVID-19 cases diagnosed during October 1, 2020–November 31, 2021, 454,010 patients and 26,462 hospitalized patients were eligible for analysis (Figure 1). Relative to patients diagnosed one day after symptom onset and adjusting for sociodemographic confounders, patients diagnosed the same day as symptom onset (adjusted odds ratio [aOR]: 1.31; 95% confidence interval [CI], 1.25, 1.37), 3–4 days after symptom onset (aOR: 1.08; 95% CI, 1.03, 1.13), and 5–21 days after symptom onset (aOR: 1.36; 95% CI, 1.31, 1.42) had increased odds of hospitalization (Table 1). Patients diagnosed the same day as their symptom onset also had higher odds of death (aOR: 1.21; 95% CI, 1.06, 1.38) (Table 1).

**Table 1.**
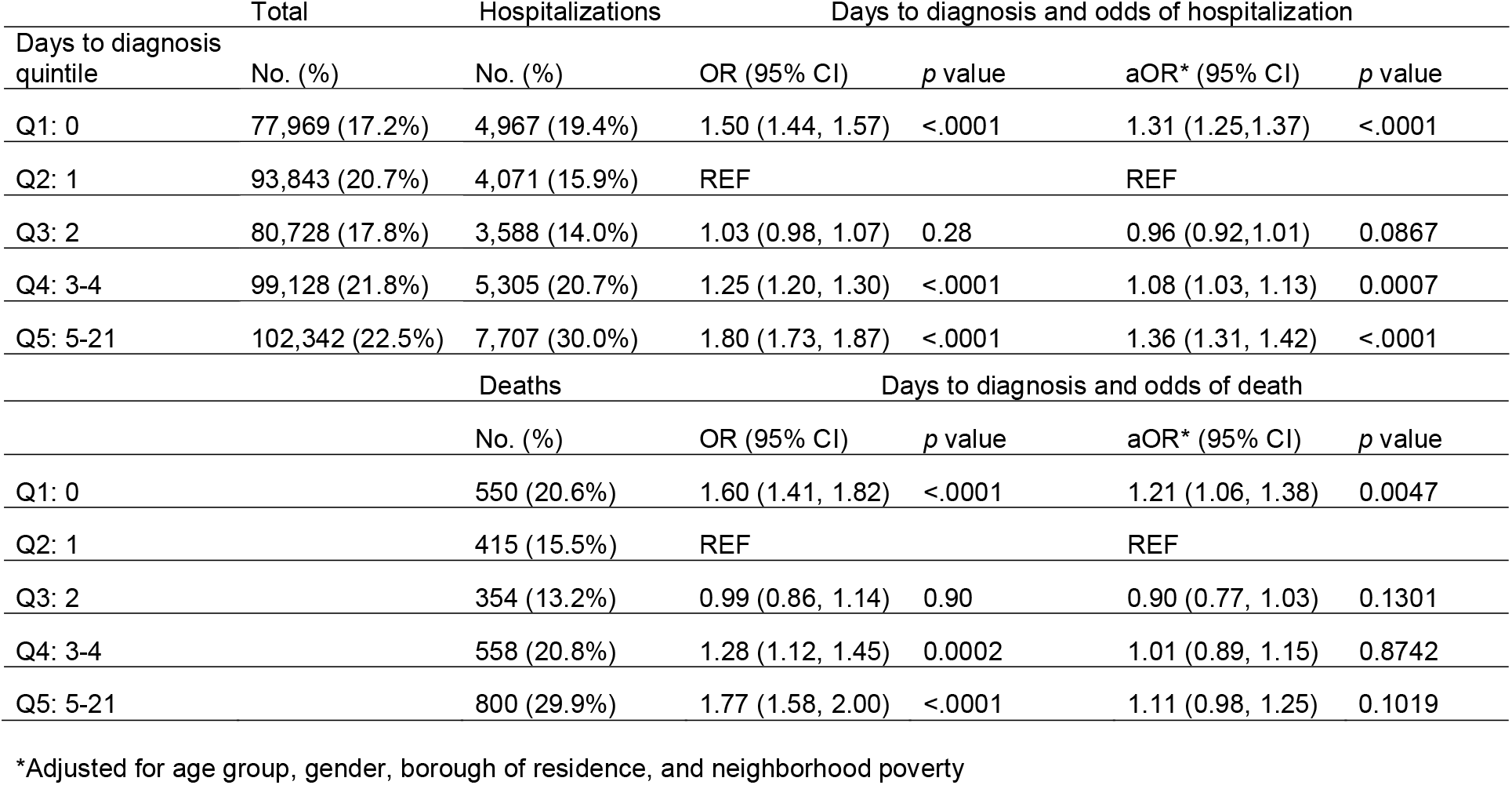
Days from COVID-19 symptom onset to diagnosis and odds of hospitalization and death, New York City, NY, USA, October 2020–November 2021

**Figure 1.**
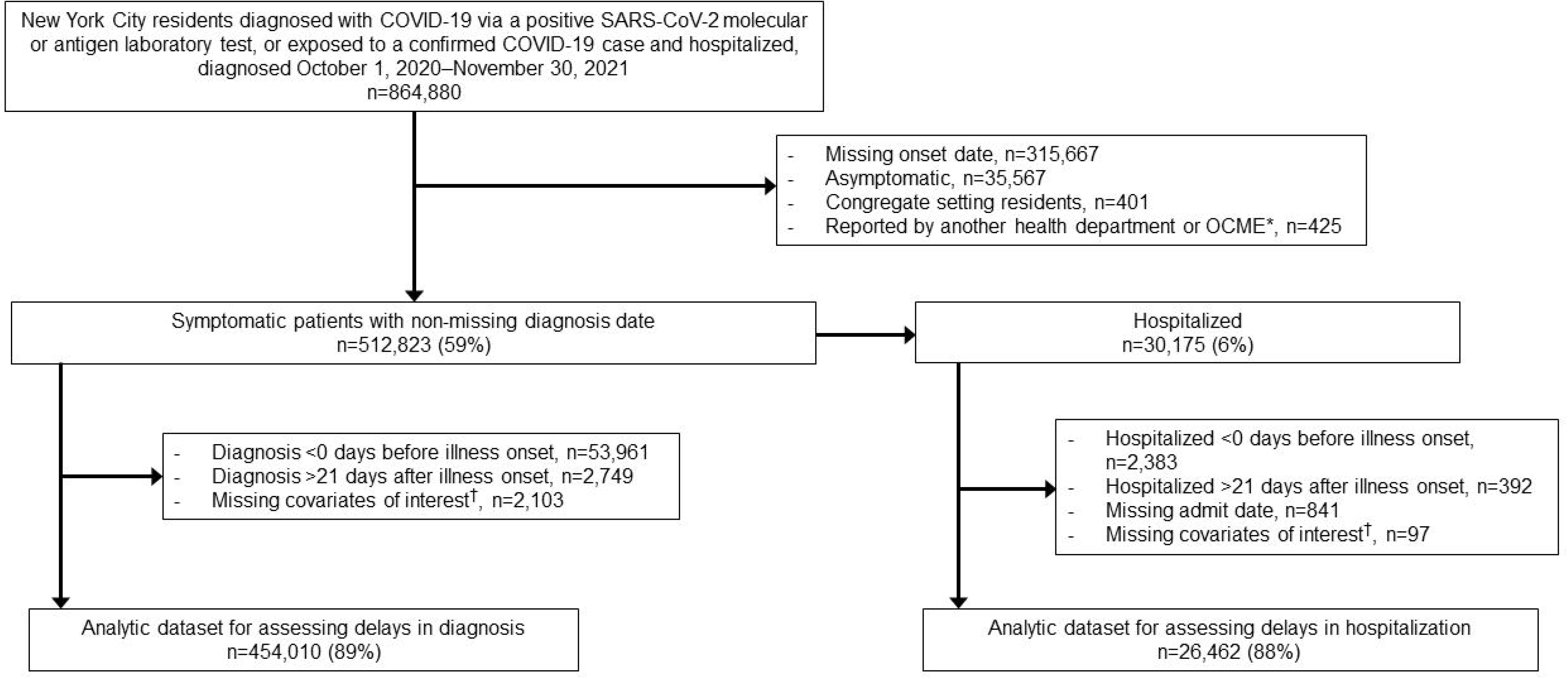
Flowchart of inclusion in analyses of delays in COVID-19 diagnosis and hospitalization, New York City, NY, USA, October 2020-November 2021 *New York City Office of Chief Medical Examiner ^†^Age group, gender, borough of residence, and neighborhood poverty

Among hospitalized patients, those hospitalized 11–21 days after symptom onset had decreased odds of death compared with patients hospitalized 0–2 days after symptom onset (aOR: 0.73; 95% CI, 0.63, 0.83) (Table 2). Patients hospitalized 6–7 days after symptom onset had increased risk of hospital stay lasting >5 days (adjusted relative risk: 1.09; 95% CI, 1.04, 1.14) (Table 2). For patients diagnosed or hospitalized 1–5 days after symptom onset, diagnosis or hospitalization during a monoclonal (mAb) antibody availability period (January 16– November 30, 2021) relative to a prior period (October 1–December 15, 2020) was not associated with improved outcomes (Appendix: Monoclonal Antibody Availability).

**Table 2.**
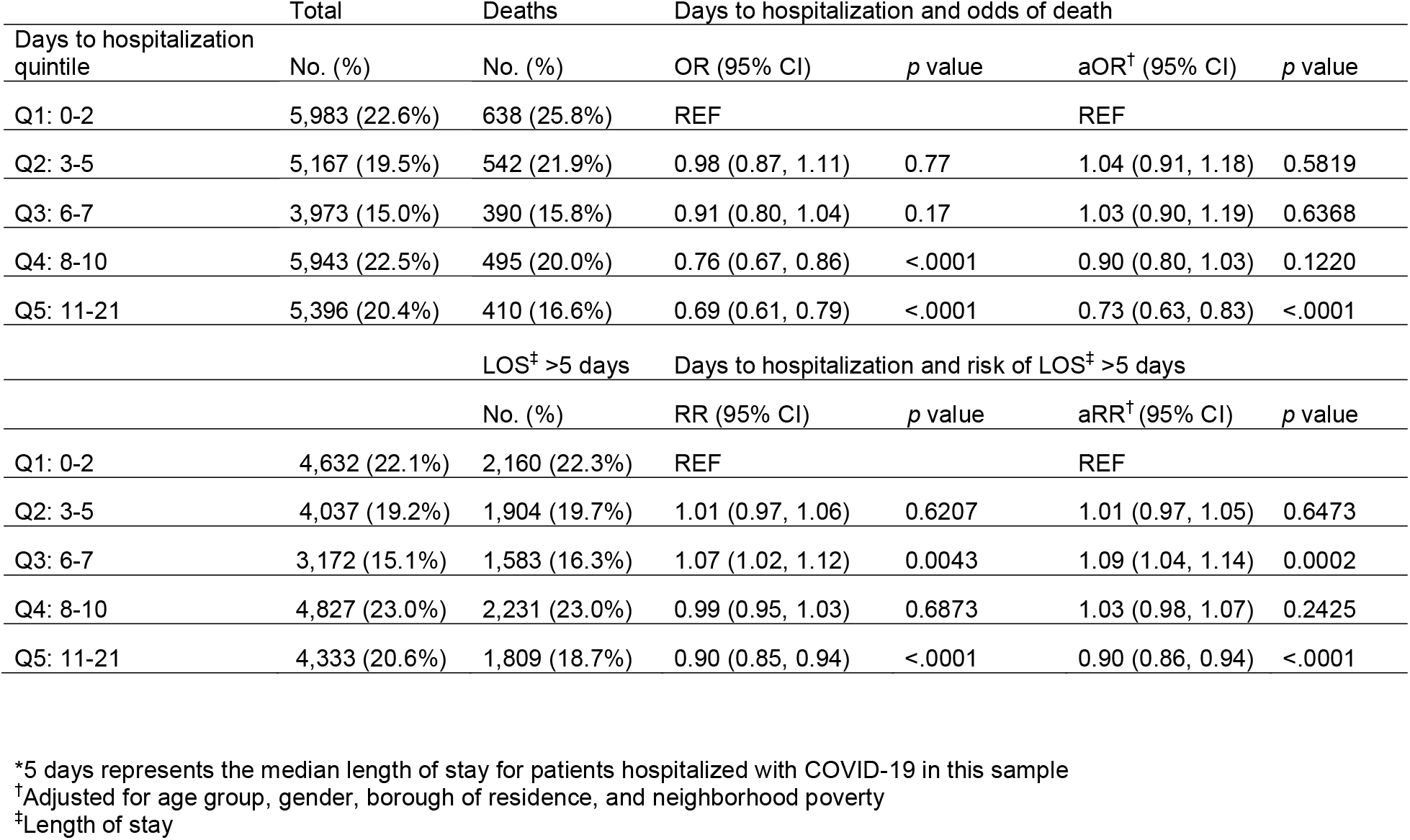
Days from COVID-19 symptom onset to hospitalization and odds of death and risk of length of stay >5 days*, New York City, NY, USA, October 2020–November 2021

We assessed whether delays from symptom onset to diagnosis varied by race/ethnicity and privilege measured by Index of Concentration at the Extremes (ICE) (*8*). Early in the study period and before Delta variant predominance (October 2020–June 2021), Black and Hispanic patients and patients in the two lowest quintiles of privilege had longer 75th percentiles for time to COVID-19 diagnosis (5 days) compared with other patients (4 days) (Figure 2). Later in the study period and during Delta variant predominance (July–November 2021), delay distributions were the same for patients of all race/ethnicities and patients in the lowest four quintiles of privilege. Among patients residing in the most privileged census tracts, the 75th percentile of time to diagnosis decreased from 4 days to 3 days (Figure 2).

**Figure 2.**
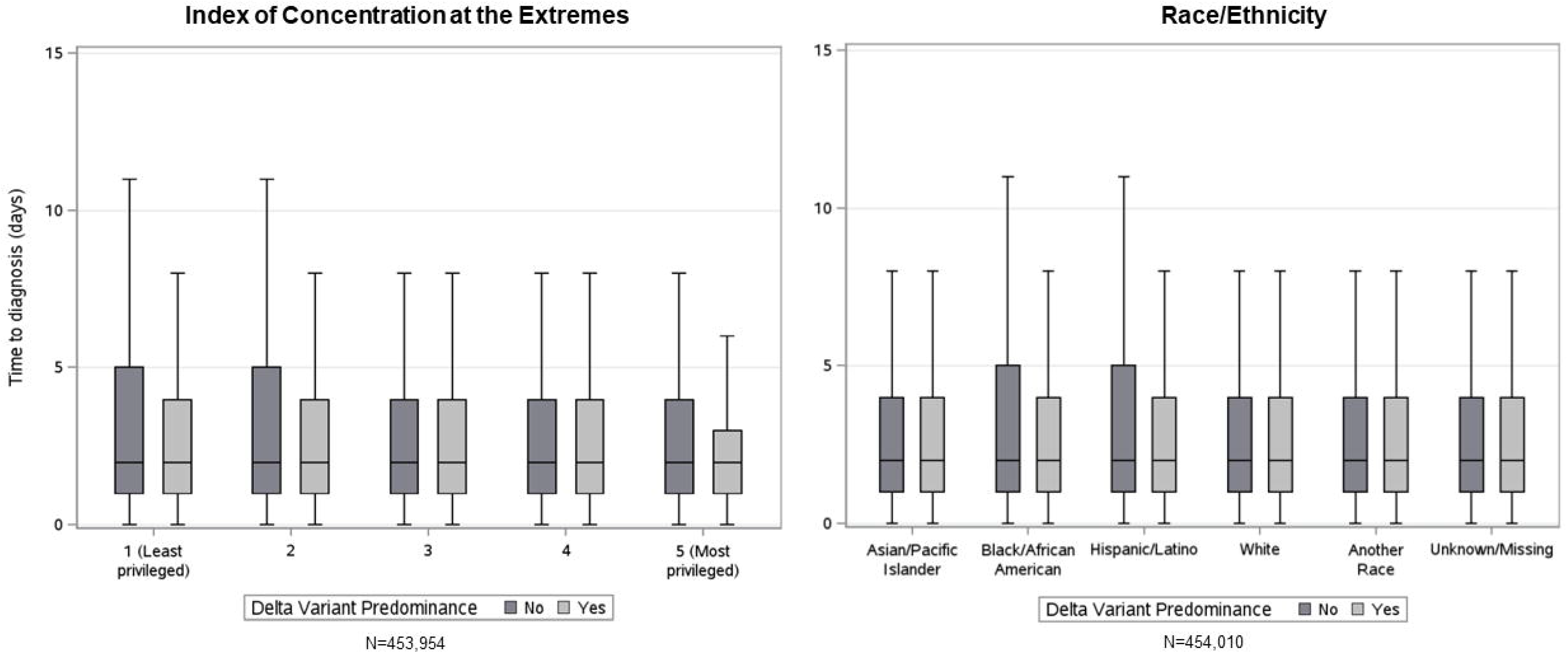
Boxplots* of delays from COVID-19 symptom onset to diagnosis by Index of Concentration at the Extremes and race/ethnicity by timing of diagnosis relative to SARS-CoV-2 Delta variant predominance,^†^ New York City, NY, USA, October 2020-November 2021 *The middle line in each box represents the median time to diagnosis. The span of each box is the interquartile range (IQR), from the 25^th^ percentile of time to diagnosis to the 75^th^ percentile. The bottom whisker is the minimum value of time to diagnosis, and the top whisker is 1.5 times the IQR Patients with time to diagnosis <0 days or >21 days were excluded. ^†^Delta variant predominance = No defined as diagnosis during October 1, 2020–June 30, 2021 Delta variant predominance = Yes defined as diagnosis during July 1, 2021–November 30, 2021.

## Conclusions

Delayed COVID-19 diagnosis ≥3 days after symptom onset was associated with increased odds of hospitalization, and hospitalization 6–7 days following symptom onset was associated with longer hospital stays, suggesting delayed testing or care-seeking might contribute to more severe outcomes. Early COVID-19 testing and diagnosis can ensure timely treatment with appropriate therapies including mAb, antivirals, and steroids, which can improve prognosis (*9, 10*). While we do not have treatment data, our findings are consistent with previous studies that support early diagnosis and hospitalization of COVID-19 patients to prevent severe outcomes (*1, 11*).

Risk of longer hospital stay was highest for patients hospitalized 6–7 days after symptom onset, consistent with the period when COVID-19 disease severity often increases (*12*). Conversely, risk of longer hospital stay and odds of death were decreased in patients hospitalized 11–21 days after symptom onset, perhaps indicating high-risk patients sought care earlier. Patients diagnosed the same day as symptom onset had greater odds of both hospitalization and death, potentially reflecting misclassification where patients were misreported as having symptom onset on their diagnosis or admission date despite earlier symptom onset.

Black and Hispanic people and residents of low-privilege areas diagnosed before Delta variant predominance experienced longer times to diagnosis, reflecting systemic racism manifested through reduced access to testing and failure of healthcare systems to build trust (*13, 14*). In late 2020, NYC Test and Trace Corps deployed mobile testing units and patient navigators to improve testing access in Task Force on Racial Inclusion and Equity (TRIE) neighborhoods, selected based on health status, social inequities, living conditions, occupation, and COVID-19 burden during wave 1 (*15*). The decrease in time to diagnosis among Black and Hispanic people and residents of the least privileged census tracts over the study period might reflect these targeted efforts to reduce testing inequities. Nevertheless, a decreased time to diagnosis was also observed among patients residing in the most privileged census tracts, indicating persistent inequities in COVID-19 testing access and resources. Sustained funding is necessary to ensure equitable access to testing and care across all communities.

Our analysis has several limitations. First, we did not have data on symptom severity and timing, which limited our ability to distinguish delays in care-seeking from slower illness progression. Second, a portion of patients with symptom onset and diagnosis on the same day could reflect recall bias or data entry errors during case investigations. Third, hospitalizations were ascertained by matching to data sources including hospital databases and health information exchanges and may be incomplete or reflect hospitalizations where SARS-CoV-2 infection was an incidental diagnosis. Nevertheless, our study adds to the limited evidence on outcomes of delays in COVID-19 care-seeking. Future studies should examine the causes of care-seeking delays and inequities to inform messaging and illuminate opportunities to direct testing and resources. In addition to vaccination, early care-seeking will be important in preventing severe COVID-19 outcomes, especially as novel therapeutics emerge.

## Data Availability

Line-level data are not publicly available in accordance with patient confidentiality and privacy laws. Publicly available data are linked below.

https://www1.nyc.gov/site/doh/covid/covid-19-data.page

## Acknowledgements

The authors thank Mary Foote, MD for her guidance and all NYC Department of Health and Mental Hygiene staff activated for the COVID-19 emergency response.

## Biographical Sketch

Laura E. Graf is a surveillance analyst in the Bureau of Hepatitis, HIV, and Sexually Transmitted Infections at the New York City Department of Health and Mental Hygiene, Queens, New York. Her interests include infectious disease epidemiology and surveillance.

## Appendix

### Statistical Analysis

Logistic regression was used to assess associations between time to diagnosis and hospitalization or death. Associations between time to hospitalization and length of stay were modeled using log-binomial regression as the outcome of length of stay >5 days was not rare (1). All models were adjusted for age group (<5, 5–14, 15–24, 25–44, 45–64, and ≥65 years-old), gender, borough of residence, and neighborhood poverty based on census tract per the American Community Survey 2015–2019 as these covariates are known to be associated with both outcome and exposure (2-4). Race/ethnicity was initially considered as an additional confounder but was excluded as a covariate from final models because of high missingness (13.0%). To assess variation in diagnosis delays, we stratified the time between symptom onset and diagnosis by race/ethnicity and privilege measured by Index of Concentration at the Extremes (ICE) (5). ICE was based on patient census tract of residence and categorized into quintiles from least privileged (highest prevalence of low-income (<$25,000), people of color households) to most privileged (highest prevalence of high-income (≥$100,000), non-Hispanic white households), per the American Community Survey 2015–2019. Of 454,010 patients in the analytical dataset, 59,151 (13.0%) had missing race/ethnicity, and 56 (0.01%) were missing a census tract and could not be assigned an ICE quintile.

Data were frozen on April 5, 2022, allowing 4 months for ascertainment of hospitalizations and deaths for patients diagnosed through the end of the study period, November 30, 2021. Only patients diagnosed starting October 1, 2020 were included, as this is when symptom onset date became reliable through case investigation. Prior to October 1, 2020, if symptom onset date was missing, it was populated by the first positive test date, and we did not want to conflate these two dates. Statistical analyses were performed using SAS Enterprise Guide, version 7.1 (SAS Institute).

### Monoclonal Antibody Availability

We further explored associations between the exposures of time to diagnosis and time to hospitalization with the outcomes of hospitalization and of death relative to monoclonal antibody (mAb) therapy availability in NYC. Patients were classified into either the pre-mAb period or the mAb availability period. Patient-level mAb data was not available, so mAb availability was considered a proxy of mAb use. mAb therapy availability in NYC increased exponentially between the weeks of December 16, 2020 and January 13, 2021 before reaching a high plateau of approximately 700 mAb treatments administered per week. mAb use decreased as cases decreased between May–July 2021, before increasing to a peak of approximately 2,000 mAb treatments administered per week during the SARS-CoV-2 Delta variant wave (New York City Department of Health and Mental Hygiene, unpub. data). Accordingly, we defined the pre-mAb therapy availability period as October 1, 2020–December 15, 2020 and the mAb availability period as January 16, 2021–November 30, 2021. Patients diagnosed during December 16, 2020– January 15, 2021 were excluded to account for delays in the rollout of mAb therapy. Only patients with a time to diagnosis of 1–5 days were included as the CDC recommends monoclonal antibodies be administered within 10 days of symptom onset, and a secondary analysis that included patients with a time to diagnosis ≤10 days yielded insignificant estimates of the odds of hospitalization and death for patients with a time to diagnosis or hospitalization >5 days.

253,926 patients met the criteria for inclusion in the mAb availability analysis. Compared with patients diagnosed 1–5 days after symptom onset in the pre-mAb availability period, odds of hospitalization were increased for patients diagnosed during the mAb availability period (aOR: 1.09, 95% CI, 1.04, 1.15). Odds of death were increased during the mAb availability period relative to the pre-mAb availability period among patients hospitalized (aOR: 1.42, 95% CI, 1.12, 1.79), but not diagnosed (aOR: 1.15, 95% CI, 0.98, 1.34), 1–5 days after symptom onset (Appendix Table).

These results contradicted our hypothesis that patients diagnosed during the mAb availability period would have decreased odds of hospitalization and death. This discrepancy could be attributed to our lack of patient-level data on mAb treatment, so diagnosis and hospitalization during the mAb availability period may not reflect mAb use. Also, more severe SARS-CoV-2 variants, including the Delta variant, emerged during the mAb availability period (6). The circulation of these variants may have resulted in increased odds of hospitalization and death in this population even with the availability of mAb. Future studies should include patient-level mAb treatment information.

---

**Appendix Table.**
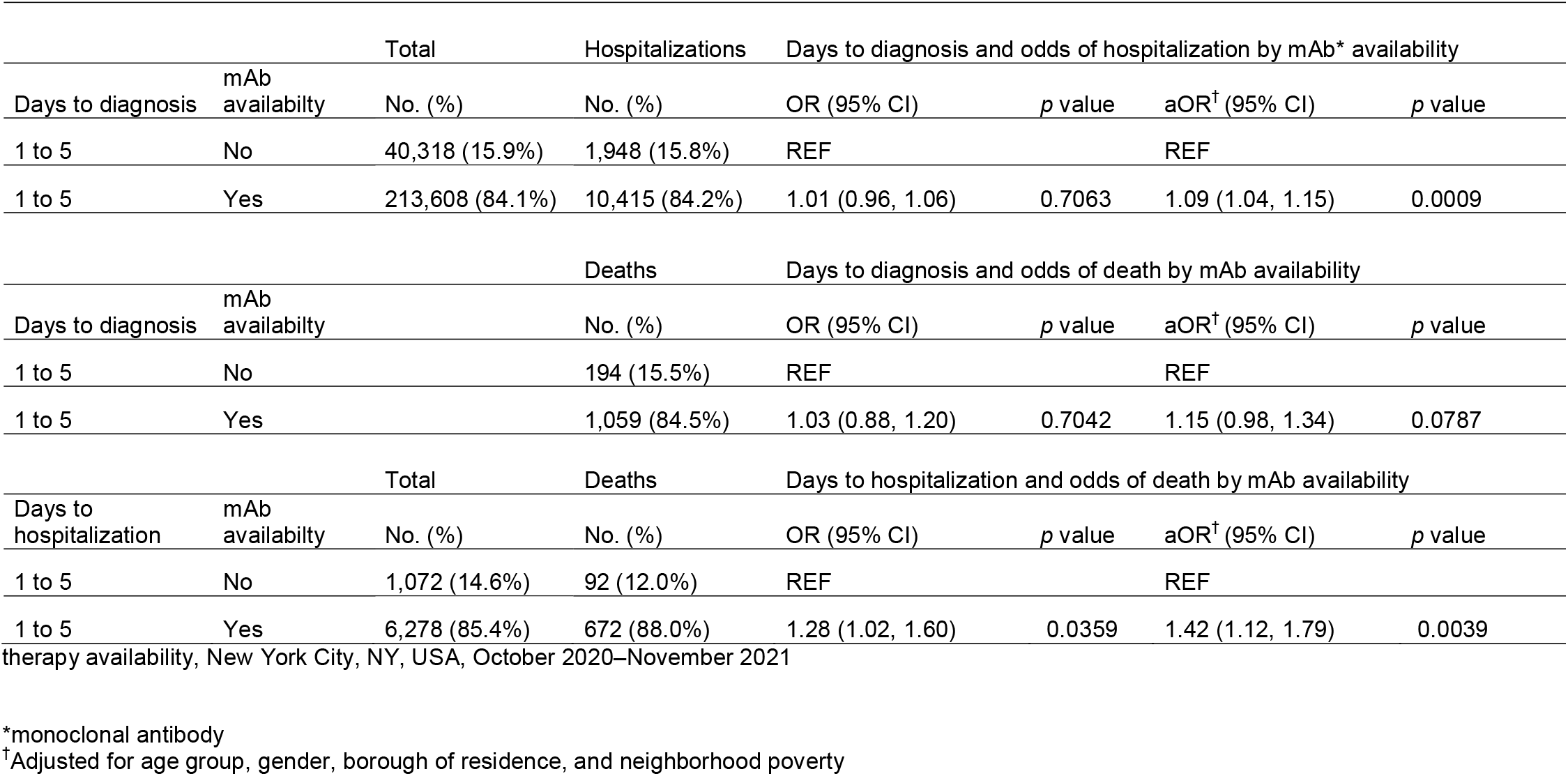
Days from symptom onset to diagnosis and hospitalization and odds of hospitalization and death by monoclonal antibody

| Days to diagnosis | mAb availability | Total | Hospitalizations | Days to diagnosis and odds of hospitalization by mAb* availability |  |  |  |
| --- | --- | --- | --- | --- | --- | --- | --- |
|  |  | No. (%) | No. (%) | OR (95% CI) | *p* value | aOR<sup>†</sup> (95% CI) | *p* value |
| 1 to 5 | No | 40,318 (15.9%) | 1,948 (15.8%) | REF |  | REF |  |
| 1 to 5 | Yes | 213,608 (84.1%) | 10,415 (84.2%) | 1.01 (0.96, 1.06) | 0.7063 | 1.09 (1.04, 1.15) | 0.0009 |
| Days to diagnosis | mAb availability | Deaths |  | Days to diagnosis and odds of death by mAb availability |  |  |  |
|  |  | No. (%) |  | OR (95% CI) | *p* value | aOR<sup>†</sup> (95% CI) | *p* value |
| 1 to 5 | No | 194 (15.5%) |  | REF |  | REF |  |
| 1 to 5 | Yes | 1,059 (84.5%) |  | 1.03 (0.88, 1.20) | 0.7042 | 1.15 (0.98, 1.34) | 0.0787 |
| Days to hospitalization | mAb availability | Total | Deaths | Days to hospitalization and odds of death by mAb availability |  |  |  |
|  |  | No. (%) | No. (%) | OR (95% CI) | *p* value | aOR<sup>†</sup> (95% CI) | *p* value |
| 1 to 5 | No | 1,072 (14.6%) | 92 (12.0%) | REF |  | REF |  |
| 1 to 5 | Yes | 6,278 (85.4%) | 672 (88.0%) | 1.28 (1.02, 1.60) | 0.0359 | 1.42 (1.12, 1.79) | 0.0039 |

therapy availability, New York City, NY, USA, October 2020–November 2021
\*monoclonal antibody
<sup>†</sup>Adjusted for age group, gender, borough of residence, and neighborhood poverty

